# LDAK-PBAT: A Pathway-Based Analysis Tool for Decoding the Genetics of Complex Diseases

**DOI:** 10.1101/2025.01.15.25320628

**Authors:** Takiy-Eddine Berrandou, Doug Speed

## Abstract

We introduce LDAK-PBAT, a novel tool for detecting gene pathways associated with complex traits. LDAK-PBAT tests pathways for significance using a heritability-based framework that controls for both the contributions of genes not in the pathway and of inter-genic SNPs.

LDAK-PBAT is computationally efficient, requiring only GWAS summary statistics and a reference panel, and when applied to human traits, can test 1000s of pathways within minutes. When assessed using simulated phenotypes, we find that LDAK-PBAT is well calibrated and has high sensitivity and specificity, outperforming both MAGMA and hypergeometric testing (F1 score of 0.734, versus 0.636 and 0.570, respectively).

When used to test 6,000 pathways for each of 37 traits (obtained from the UK Biobank, Millions Veterans Program and psychiatric genomics consortium; sample sizes ranging from 17,014 to 1,114,458), LDAK-PBAT identified 4,861 significant pathways (P < 0.05/6000), which is substantially more than found by MAGMA (97) and hypergeometric testing (522).

LDAK-PBAT emerges as a powerful tool for advancing genetic research, offering advanced insights into complex diseases. Its precise estimation of pathway enrichment and heritability, along with superior performance in detecting significant pathways. LDAK-PBAT holds significant promise for uncovering the genetic underpinnings of complex diseases and driving forward the development of personalized medicine.

## I. Introduction

Genome-wide association studies (GWAS) have significantly advanced our understanding of the genetic basis of complex diseases, providing insights into disease mechanisms and potential therapeutic targets [1]. Traditionally, GWAS analyses test individual single nucleotide polymorphisms (SNPs) for association with phenotypes[2]. However, it is increasingly common to perform pathway-based analyses [3], which evaluate sets of SNPs defined by their involvement in biological pathways. This approach can be more powerful than single-SNP analyses as it considers the collective effect of functionally related genes, enhancing the ability to detect associations that may be missed when examining SNPs individually. Further, it can be more biologically meaningful, because the significant pathways provide insight into the functional mechanisms underlying the trait.

We introduce LDAK-PBAT, a novel tool for pathway-based genetic analysis that employs a heritability enrichment framework [4]. For each pathway, LDAK-PBAT estimates the heritability contributed by SNPs in genes within the pathway, then compares this to the estimated heritability of SNPs in genes outside the pathway (while also allowing for the contribution of inter-genic SNPs). LDAK-PBAT returns the estimated heritability enrichment of the pathway, and a corresponding p-value. This approach ensures that the cumulative effect of all genes within a pathway is considered, enhancing the detection power for pathway-level associations.

In this study, we validate the performance of LDAK-PBAT using extensive simulations and real data from the UK Biobank (UKBB) [5], the Million Veteran Program (MVP) [6], and the Psychiatric Genomics Consortium (PGC) [7]. Our simulations assess the tool’s accuracy and robustness by generating continuous and binary phenotypes across various genetic architectures. Additionally, we performed comparative analyses with established gene-set methods such as Multi-marker Analysis of GenoMic Annotation (MAGMA) [8] and hypergeometric testing [9], demonstrating LDAK-PBAT’s superior sensitivity and precision in detecting significant pathways.

LDAK-PBAT is freely available within our software package LDAK, which includes full documentation, test data, and example scripts (see web resources for a link to the LDAK website).

## II. Methods

### 1. Overview of LDAK-PBAT

LDAK-PBAT extends SumHer [4], our existing tool for heritability analysis using GWAS summary statistics. Suppose the set S indexes the SNPs within the pathway being tested, while the set G indexes all genic SNPs (i.e., S is a subset of G). Further, let E[h²_j_] denote the expected heritability of SNP j, while I_S(j_ and I_G(j_ indicate whether the SNP is within S and G, respectively. LDAK-PBAT estimates the importance of the pathway using the following (genome-wide) heritability model

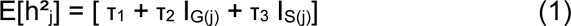

where τ_1_, τ_2_ and τ_3_ measure, respectively, the average heritability contribution of all SNPs, of genic SNPs and of SNPs in the pathway. LDAK-PBAT tests whether τ_3_ is significantly greater than zero, which indicates that the average per-SNP heritability for genes in the pathway is significantly than that for genes not in the pathway. This LDAK-PBAT uses a competitive testing framework [10], where the genetic contribution of the pathway is compared to a genome-wide background, ensuring that the analysis accounts for the collective effects of all genes both inside and outside the pathway.

LDAK-PBAT solves Equation 1 by constructing an approximate likelihood of GWAS test statistics given the heritability model, then finding τ_1_, τ_2_ and τ_3_ that maximize this likelihood. It tests the significance of τ_3_ by comparing the full likelihood to the reduced likelihood obtained by setting τ_3_ =0. Note that while it is possible to run LDAK-PBAT using the original version of SumHer, this would require a separate analysis for each pathway (and would generate thousands of redundant output files). By contrast, LDAK-PBAT is able to test all pathways within a single analysis, and stores only the key statistics corresponding to each pathway (i.e. its estimated heritability enrichment and significance).

#### Workflow of LDAK-PBAT

LDAK-PBAT employs a one-step methodology to analyze pathway associations directly. Unlike methods such as MAGMA, which first compute gene-level associations and then aggregate these to the pathway level, LDAK-PBAT estimates pathway heritability, enrichment, and significance (p-value) in a single step. While pathways are analyzed one at a time, this direct approach is highly efficient, allowing the tool to test thousands of pathways within minutes using only GWAS summary statistics.

LDAK-PBAT’s workflow integrates GWAS summary statistics and a pre-computed tagging file containing SNP lists, pathway definitions, and linkage disequilibrium (LD) patterns. These inputs feed into the heritability model to estimate pathway contributions to trait variance by optimizing τ_1_, τ_2_ and τ_3_. The outputs include pathway-level heritability enrichment estimates and p-values, enabling insights into the genetic architecture of complex traits. An overview of this process is illustrated in Figure 1.

**Figure 1.**
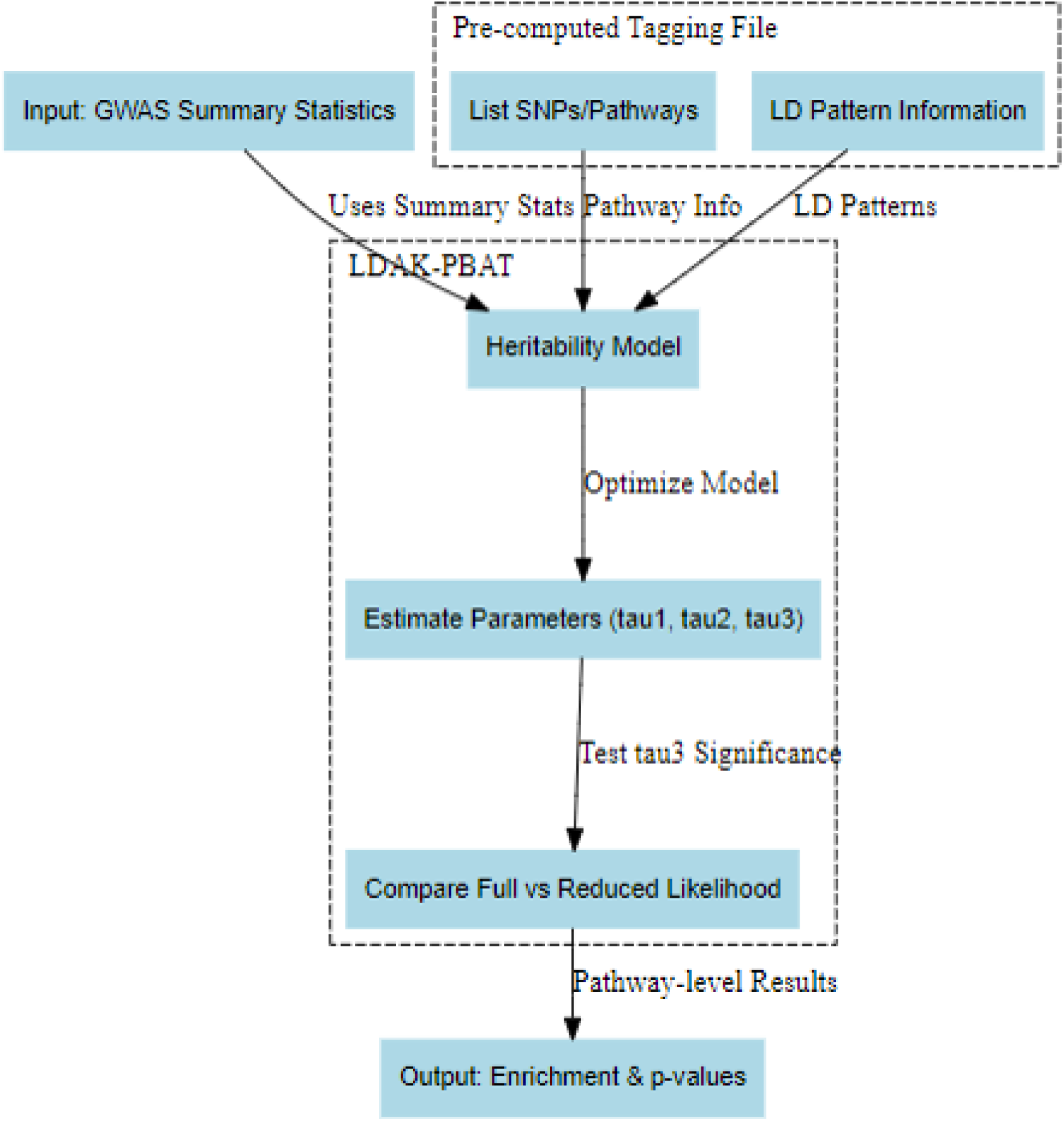
Overview of the LDAK-PBAT workflow. The analysis begins with two main inputs: (1) GWAS summary statistics, and (2) a pre-computed tagging file that includes SNP lists/pathways and linkage disequilibrium (LD) pattern information. The pre-computed tagging file facilitates efficient incorporation of pathway-specific genetic information. The heritability model, E[h²_j_] = [ τ_1_ + τ_2_ I_G(j)_ + τ_3_ I_S(j)_], integrates these inputs to estimate pathway contributions to trait variance by optimizing key parameters (τ_1_, τ_2_, τ_3_). The workflow then evaluates the significance of τ_3_ using a likelihood comparison (full vs reduced model). The outputs include pathway-level heritability enrichment estimates and associated p-values, providing insights into the genetic architecture of complex traits.

### 2. UK Biobank (UKBB), Million Veteran Program (MVP), and Psychiatric Genomics Consortium (PGC) Summary Statistics

We accessed UKBB [5] data through application 21,432 and computed summary statistics for ten key phenotypes, including body mass index, height, hypertension, neuroticism score, and systolic blood pressure. Summary statistics were derived using classical linear regression on a cohort of 50,000 unrelated, White British individuals, incorporating 13 covariates (age, sex, Townsend Deprivation Index, and ten principal components) to minimize confounding and population stratification. Single-SNP analysis was performed on up to 200,000 individuals.

For the MVP [6] and PGC [7] phenotypes, we used summary statistics from previously published GWASs. The MVP data included 18 phenotypes such as type 2 diabetes, cardiovascular diseases, number of cigarettes per day, lipid levels, and blood pressure, with average sample sizes of 242,207 (ranging from 17,014 to 1,114,458). The PGC data covered nine phenotypes, including attention-deficit hyperactivity disorder, Alzheimer’s disease, alcohol use, autism spectrum disorder, bipolar disorder, eating disorders, major depressive disorder, post-traumatic stress disorder, and schizophrenia, with average sample sizes of 256,250 (ranging from 46,351 to 762,917).

### 3. Pathway Definitions

We selected 6000 pathways from the MSigDB (v7.5.1) database [11-13]. These pathways were chosen based on specific criteria, including:

- Number of SNPs (N_SNP): Pathways with SNP counts ranging from 5,000 to 70,000 were included.
- Number of Genes (N_Genes): Pathways containing between 5 and 1,078 genes were selected. The pathways were drawn from various collections within MSigDB, including: BIOCARTA: 52, Kyoto Encyclopedia of Genes and Genomes (KEGG): 104, Pathway Interaction Database (PID): 128, Gene Ontology Cellular Component (GOCC): 294, Gene Ontology Molecular Function (GOMF): 440, Gene Ontology Biological Process ( GOBP): 2,479, WikiPathways (WP): 272, REACTOME: 536 and Other (published): 1,695.

### 4. Simulation of null phenotypes to Assess Type I Error Calibration

To assess Type I error under the null hypothesis (no true pathway enrichment), we first randomly selected 50,000 individuals from the UK Biobank dataset, ensuring each had genotype data for the relevant SNP set. We then permuted their phenotype values 100 times to produce 100 simulated null traits, thus preserving the original genotype structure while breaking any true genotype–phenotype association. For each permuted trait, we ran a standard genome-wide association study (GWAS) to obtain summary statistics. Next, we applied LDAK-PBATto_Stat2_, which captures the deviation of each pathway’s absolute heritability from its expected contribution, as recommended in previous work [14]. A pathway was declared “significant” if its Z_Stat2_ exceeded the corresponding one-sided Z-threshold for various nominal *p*-value cutoffs (0.05, 10⁻³, 10⁻⁵, and 0.05/6000—e.g., *Z* > 4.2649 for *p* < 10⁻⁵). Counting the number of significant pathways across all 100 null replicates allowed us to estimate the empirical Type I error rate at each threshold.

### 5. Simulation to heritable phenotypes Evaluate LDAK-PBAT Accuracy and Robustness

We generate 1125 continuous phenotypes and 2250 binary phenotypes. For the continuous phenotypes, we varied the heritability (h²) values (0.1, 0.3, or0.5), the number of causal SNPs (2000, 5000, and 10000), and the enrichment weights for different annotations (five values, defined below), resulting in 45 different scenarios. For binary phenotypes, we also varied the prevalence rates at 0.05 and 0.1, resulting in 90 different scenarios.

We sampled 20 pathways of varying sizes from the msigDB database, ranging from a minimum of 6,000 SNPs to a maximum of 50,000 SNPs, covering 5 to 1078 genes. For the annotation enrichment, we adjusted the probability weights to include causal variants as follows: genic SNPs inside the 20 selected pathways were assigned weights of x2, x5, x10, x20, and x50, while genic SNPs outside these pathways were given a weight of x2, and intergenic SNPs were assigned a weight of x1. This variation in enrichment probabilities allowed us to simulate diverse levels of genetic influence within and outside of pathways.

To ensure robustness and reproducibility, we generated 25 replicates for each scenario defined by the combination of heritability, number of causal variants, and annotation enrichment weights. We used genotyping data from 50,000 individuals from the UK Biobank (UKBB) to generate the phenotypes. Once the phenotypes were generated, we conducted genome-wide association studies (GWAS) to produce summary statistics, focusing on the HapMap SNPs (∼1.1 million SNPs) [15]. This setup allowed us to create realistic scenarios reflecting the complexity of genetic data.

The simulated phenotypes, pathway sizes, and SNP configurations were designed to mimic real-world genetic architectures. By varying the probability that an annotation includes a causal variant, we could evaluate the performance of LDAK-PBAT under different conditions of pathway enrichment and genetic influence. These simulations provided a robust framework for validating the accuracy and precision of LDAK-PBAT in estimating pathway heritability and detecting significant pathways. The diverse scenarios and extensive replicates ensured that the tool’s performance was thoroughly tested, demonstrating its reliability in handling complex genetic datasets.

### 6. Evaluation of pathway heritability and heritability enrichment

We employed a range of statistical methods to rigorously analyse the data and validate the performance of LDAK-PBAT. For the evaluation of pathway heritability and heritability enrichment, we used root mean square error (RMSE) and correlation coefficients as key metrics. RMSE was calculated to measure the average deviation of the estimated heritability from the true heritability values, while correlation coefficients were used to assess the linear relationship between the estimated and true values.

### 7. Evaluation Criteria for Comparative Analysis

To comprehensively assess the performance of LDAK-PBAT in comparison with other gene-set methods, we used key metrics: sensitivity, precision, F1 score, and specificity. These metrics were calculated based on simulations where positive (i.e., causal) pathways were defined using their true enrichment values, determined by the simulation parameters. Specifically, true heritability enrichment was calculated by summing the heritability of causal variants within each pathway and comparing it to the expected enrichment given the size of the pathway. True hypergeometric enrichment was calculated by counting the number of causal SNPs within each pathway and comparing it to the total number of SNPs inside and outside the pathway. A pathway was considered a true causal pathway if both its true heritability enrichment and true hypergeometric enrichment met or exceeded a predefined threshold. To ensure robustness in identifying true causal pathways, we considered a series of thresholds for "true" pathway enrichment, ranging from 1.1 to 10 in increments of 0.1. For each threshold, we evaluated whether a pathway met the criteria to be considered causal. This allowed us to define a set of true causal pathways for each threshold.

Once the true causal pathways were defined at various thresholds, we analysed each pathway using the LDAK-PBAT, MAGMA, and Hypergeometric methods. For each method and threshold combination, we identified the pathways predicted to be significant and calculated performance metrics: True Positives (TP), False Positives (FP), False Negatives (FN), and True Negatives (TN). From these values, we derived the following metrics:

*-* Sensitivity (also known as recall) measures the proportion of true positive results correctly identified by the method: Sensitivity = True Positives / (True Positives + False Negatives).
*-* Precision (also known as positive predictive value) indicates the proportion of positive results that are true positives: Precision = True Positives / (True Positives + False Positives).
*-* F1 Score is the harmonic mean of precision and sensitivity, balancing both concerns: F1 Score = 2 × (Precision × Sensitivity) / (Precision + Sensitivity).
*-* Specificity measures the proportion of true negative results correctly identified by the method: Specificity = True Negatives / (True Negatives + False Positives).

Finally, we computed the average values of these metrics across all thresholds and scenarios, providing a comprehensive measure of each method’s performance in identifying true causal pathways. These averaged metrics were then used to compare the effectiveness of LDAK-PBAT, MAGMA, and Hypergeometric testing across different P-value thresholds.

### 8. Existing Gene-set Methods for Comparative Analysis

For the comparative analysis, we employed two widely recognized gene-set methods: hypergeometric testing and MAGMA. Both methods are designed to support the competitive testing framework, which evaluates whether the association of genes within a pathway is significantly greater than the association of genes outside the pathway.

Hypergeometric testing is a widely used method for pathway enrichment analysis, leveraging overlap statistics between significant genetic elements and pathway components. It evaluates the null hypothesis that genetic elements (e.g., SNPs or genes) within a pathway are no more associated with the trait than elements outside the pathway. A contingency table is used to calculate overlaps, comparing significant and non-significant elements inside and outside the pathway.

For this analysis, we employed four variants of hypergeometric enrichment testing, tailored to different definitions of pathway components and genetic significance:

a. Hypergeometric Enrichment Based on Significant Clumped SNPs (Hyper_SNP_Hits): This approach calculates enrichment using the count of GWAS-significant clumped SNPs. For each pathway, the count of significant SNPs within the pathway (N_sig SNP in pathway_) is compared against the total number of SNPs in the pathway (N_SNP in pathway_) and genome-wide SNP counts (N_total SNP_). So, the null hypothesis is "The significant SNPs in the pathway are proportional to their occurrence genome-wide."
b. Hypergeometric Enrichment Based on Genes Harboring Significant SNPs (Hyper_Gene_GWAS_Hits): This method evaluates pathway enrichment by counting genes harboring GWAS-significant SNPs. A gene is considered significant if it contains at least one SNP with a p-value below the genome-wide significance threshold (5×10^−8^). The pathway-specific count of these genes (N_genes with sig SNP in pathway_) is compared to the total number of genes in the pathway (N_genes in pathway_) and genome-wide gene counts (N_total genes_). Therefore, the null hypothesis is "The genes with significant SNPs are not more enriched in the pathway than expected by chance."
c. Hypergeometric Enrichment Based on Genes with SNPs Passing Bonferroni Gene Threshold (Hyper_Gene_MinP_Hits): This variant focuses on genes with at least one SNP whose minimum p-value (MinP) passes a Bonferroni-corrected threshold based on the total number of genes (0.05/N_total genes_). The count of these significant genes in the pathway (N_genes with SNP MinP in pathway_) is compared to pathway and genome-wide gene counts. The null hypothesis is "Genes with highly significant SNPs, as measured by MinP, are not overrepresented in the pathway."
d. Hypergeometric Enrichment Based on Significant Genes from GBAT Analysis (Hyper_Gene_GBAT_Hits): This method builds on gene-based association testing (GBAT), which has shown superior performance over other gene-based approaches. A gene is considered significant if its GBAT p-value is below a Bonferroni threshold based on the total number of genes (0.05/N_total genes_). Pathway enrichment is calculated based on the count of significant genes in the pathway (N_GBAT sig genes in pathway_), pathway-wide gene counts, and genome-wide gene counts. So, the null hypothesis is "Significant genes identified by GBAT are not enriched in the pathway." Multi-marker Analysis of GenoMic Annotation (MAGMA) [8] employs a linear regression approach to evaluate pathway enrichment. Gene-level statistics are regressed on pathway definitions, with the slope of the regression indicating the degree of enrichment. This method allows for straightforward derivation of p-value significance, facilitating the assessment of pathway-level associations.

The selection of hypergeometric testing and MAGMA for comparative analysis is based on their widespread use and established reliability in the field of genetic pathway analysis. Both methods are well-documented and have been extensively validated in numerous studies, making them suitable benchmarks for evaluating the performance of LDAK-PBAT. Hypergeometric testing provides a straightforward and intuitive approach to assessing pathway enrichment by focusing on the overlap between significant genes and pathway genes. This method is particularly useful for its simplicity and ease of interpretation. MAGMA, on the other hand, offers a more sophisticated statistical approach through linear regression, allowing for a nuanced analysis of gene-level data within the context of pathway definitions. Its capability to handle large-scale genetic data and derive precise p-values makes it a robust tool for pathway analysis.

In the comparative studies, both hypergeometric testing and MAGMA were applied to the same datasets used for LDAK-PBAT to ensure consistency in the evaluation.

### 9. Sensitivity Analysis of LDAK-PBAT

#### Impact of SNP List and Reference Panel Variations

To evaluate the robustness of LDAK-PBAT against variations in SNP lists and reference panels, we conducted a sensitivity analysis on a subset of 225 simulated phenotypes, restricting the analysis to 100 pathways.

For the SNP list variation analysis, we compared models using the same UK Biobank individuals (n=2500) as the reference panel but with different SNP lists: one based on 1 million HapMap SNPs and the other utilizing an expanded list of 7.1 million SNPs. This allowed us to assess the consistency of heritability enrichment and p-value significance between the two SNP lists.

For the reference panel variation analysis, we held the SNP list constant at 1 million HapMap SNPs and compared the analyses using UK Biobank individuals (n=2500) versus 404 non-Finnish European individuals from the 1000 Genomes Project (1000G) [15]. This comparison aimed to determine the impact of different reference populations on heritability enrichment and p-value significance.

#### Impact of Genic Component Exclusion

To evaluate the robustness of LDAK-PBAT when excluding the genic annotation component, we tested a modified heritability model (Equation 1) comprising only τ_1_ (overall heritability contribution of all SNPs) and τ_3_ (heritable contribution of SNPs within pathways), omitting τ_2_ (the genic SNPs component). This analysis was conducted using GWAS summary statistics for 37 traits from the UK Biobank (n = 10), PGC (n = 9), and MVP (n = 18) datasets, encompassing diverse phenotypic and genetic architectures.

For each trait, we:

1. Applied LDAK-PBAT to evaluate 6000 predefined pathways using the full heritability model with the genic component included.
2. Repeated the analysis with the modified model excluding the genic component (τ_2_).
3. Compared pathway-level Z-statistics (Z_Stat2_) between the two models at different thresholds (e.g., p=0.05/6000, corresponding to Z>4.3054).

We identified potential outliers, defined as pathways where the ratio of Z_Stat2_ between the two models exceeded 3 or fell below 1/3. Outliers were flagged as pathways with highly discordant enrichment results when the genic component was excluded.

#### Impact of Adding an Intercept

We extended the heritability model (Equation 1) by including an intercept term, analogous to LD Score Regression, to account for confounding due to population structure and relatedness. This intercept term is designed to capture uniform inflation in test statistics across all pathways. To evaluate the impact of adding the intercept, we conducted the analysis using GWAS summary statistics for 37 traits from the UK Biobank (n = 10), PGC (n = 9), and MVP (n = 18) datasets. For each trait:

1. LDAK-PBAT was applied to evaluate 6000 predefined pathways using the default heritability model without the intercept.
2. The analysis was repeated with the intercept included.
3. Pathway-level Z_Stat2_ values were compared between the two models at different thresholds (e.g., p=0.05/6000, corresponding to Z>4.3054).

Pathways were categorized as concordant or discordant based on whether they were declared significant under both models, and outliers were identified as pathways with highly discordant Z_Stat2_ ratios (e.g., exceeding a factor of 3 or below 1/3).

## III. Results

### LDAK-PBAT is well calibrated

Table 1 summarizes the Type I error outcomes from 100 null (permuted) phenotypes at four nominal *p*-value thresholds—0.05, 10⁻³, 10⁻⁵, and 0.05/6000—using Z_Stat2_. Notably, at *p* = 10⁻⁵ and the Bonferroni level (0.05/6000), zero pathways were declared significant across all simulations, in line with the nominal expectation of approximately 6 and 5 false positives, respectively, under a naive binomial model. Even at the more permissive thresholds, the observed number of hits (e.g., 10,774 at *p* = 0.05) remained below the naive expectation of 30,000, indicating a conservative (i.e., deflated) behavior rather than inflation. The final column shows the binomial *p*-value for obtaining at least as many observed hits under an independence assumption; because the observed was fewer than expected at most thresholds, these upper-tail *p*-values are effectively 1.00. Overall, these results confirm that Z_Stat2_ is well-calibrated and shows no sign of inflation at stringent *p*-value cutoffs, thus providing robust Type I error control.

**Table 1.** Observed vs. expected significant pathways for **Z_Stat2_** under the null (100 permuted phenotypes) at four nominal *p*-value thresholds.

| Threshold | p | Z-threshold | Observed Hits ($Z_{Stat2}$) | Naive Expected | Binomial p |
| --- | --- | --- | --- | --- | --- |
| 1 | 0.05 | 1.6449 | 10,774 | 30,000 | 1 |
| 2 | $10^{-3}$ | 3.0902 | 4 | 600 | 1 |
| 3 | $10^{-5}$ | 4.2649 | 0 | 6 | 1 |
| 4 | 0.05/6000 | 4.3054 | 0 | 5 | 1 |

### Accuracy of heritability estimation in pathway analysis by LDAK-PBAT: evidence from simulation studies

LDAK-PBAT’s estimation accuracy for pathway heritability and heritability enrichment has been rigorously validated through extensive simulations, with the results displayed in Figures 2 and 3. These figures together illustrate the tool’s robust performance across a spectrum of genetic complexities involving 1,125 simulated phenotypes and 20 distinct pathways of varying sizes.

**Figure 2.**
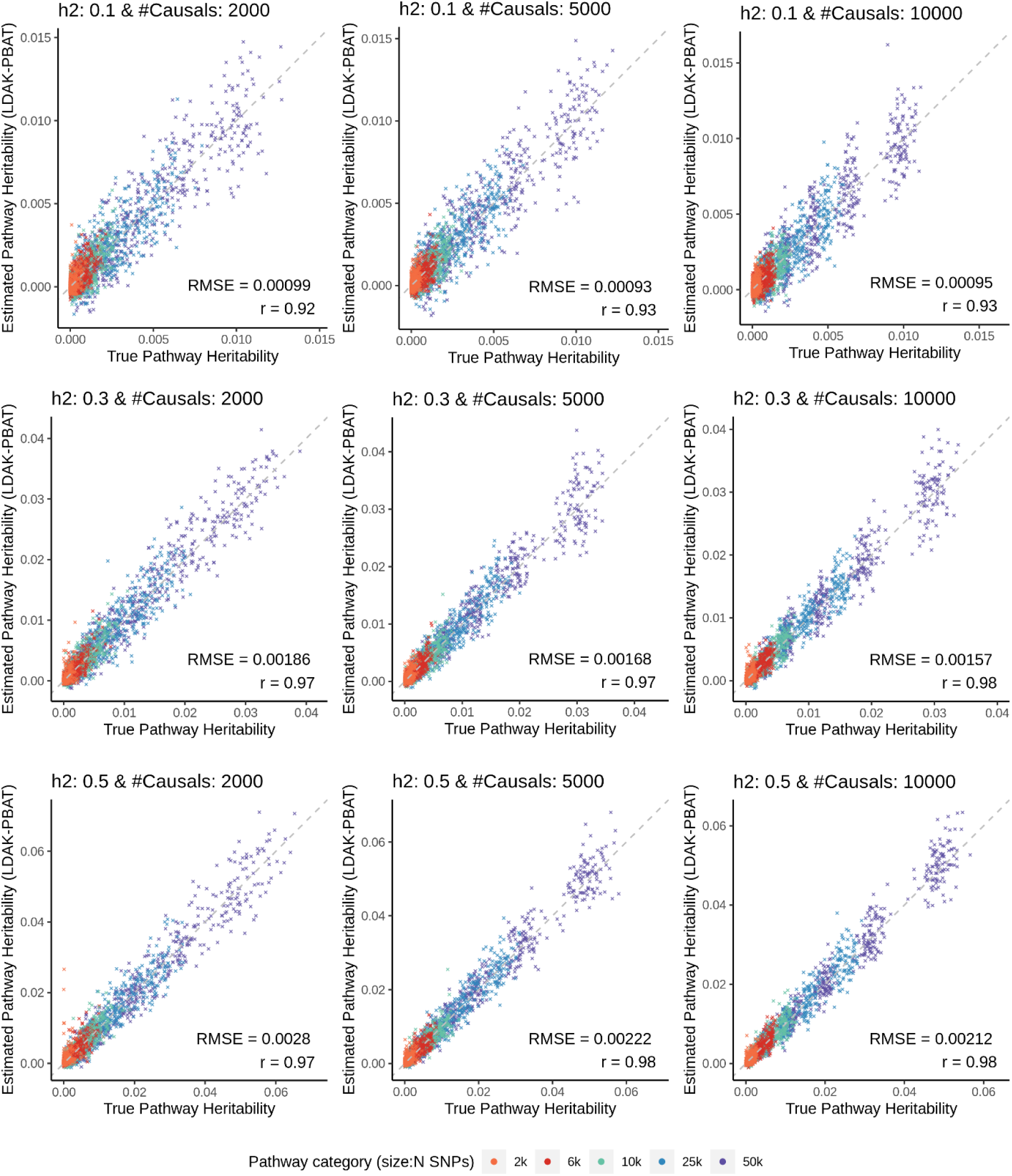
Assessment of LDAK-PBAT Performance in Pathway Heritability Estimation Across Multiple Genetic Architectures. The figure presents a comparative analysis of estimated pathway heritability versus true pathway heritability for simulated phenotypes. The scatter plots are organized into a matrix of panels, each representing a unique scenario based on varying levels of heritability (h² values of 0.1, 0.3, 0.5) and numbers of causal SNPs (2000, 5000, 10000). Within each panel, the x-axis denotes the true pathway heritability, while the y-axis represents the heritability estimated by LDAK-PBAT. Data points are color-coded to reflect the size of the pathway category, ranging from 2k SNPs (orange) to 50k SNPs (dark blue). The alignment of data points with the diagonal line in each panel indicates the degree of accuracy in heritability estimation by LDAK-PBAT.

**Figure 3.**
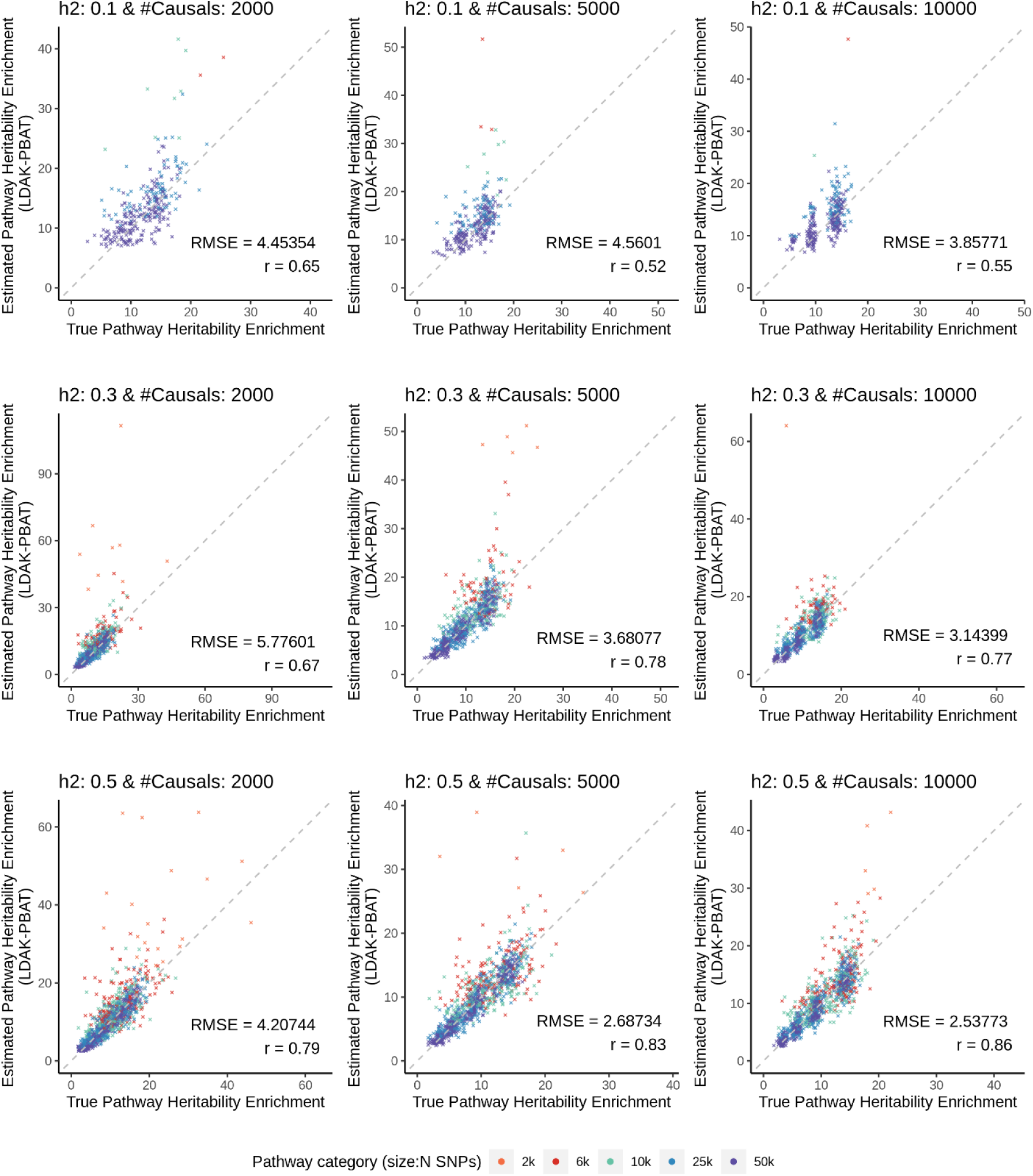
Comparison of Significant Estimated Pathway Heritability Enrichment to True Pathway Heritability Enrichment Across Diverse Genetic Configurations. The figure depicts the estimated pathway heritability enrichment against the true pathway heritability enrichment for a range of genetic architectures. The figure is organized into a grid of nine scatter plots, each representing a different combination of heritability (h² values of 0.1, 0.3, and 0.5) and numbers of causal SNPs (2000, 5000, and 10000). The x-axis represents the true pathway heritability enrichment, while the y-axis shows the estimated enrichment as determined by LDAK-PBAT. Data points are color-coded to reflect the size of the pathway category, ranging from 2k SNPs (orange) to 50k SNPs (dark blue). The proximity of points to the diagonal line across the scatter plots signals the accuracy of LDAK-PBAT’s estimations.

For pathway heritability, as demonstrated in Figure 2, LDAK-PBAT exhibited exceptional precision at a lower heritability level (h² = 0.1) with RMSE values of 0.00099, 0.00093, and 0.00095 for sets of 2000, 5000, and 10000 causal SNPs, respectively, and corresponding correlation coefficients of 0.92 across all three SNP sets. This precision persisted as heritability increased to 0.3, evidenced by RMSEs of 0.00186, 0.00168, and 0.00157 and correlation coefficients of 0.97. At the highest heritability level tested (h² = 0.5), the tool achieved RMSE values of 0.00280, 0.00222, and 0.00212, along with correlation coefficients of 0.97, 0.98, and 0.98 for the respective SNP causality sets, highlighting its capability to deliver accurate estimations in scenarios with greater genetic diversity.

Turning to pathway heritability enrichment, LDAK-PBAT demonstrated consistent performance across all configurations. At the lowest heritability level (h²=0.1), the RMSE values ranged from 4.45 (for 2000 causal SNPs) to 3.86 (for 10000 causal SNPs), and the correlation coefficients were 0.65 and 0.55, respectively. As heritability increased to h²=0.3, the RMSE values decreased, ranging from 5.78 (for 2000 causal SNPs) to 3.14 (for 10000 causal SNPs), with correlation coefficients improving to a range of 0.67–0.77. For the highest heritability (h²=0.5), the RMSE values were the lowest, ranging from 4.21 (for 2000 causal SNPs) to 2.54 (for 10000 causal SNPs), and the correlation coefficients peaked at 0.86, indicating a strong linear relationship between the true and estimated enrichment values.

These results underscore the precision of LDAK-PBAT’s estimations in both heritability and enrichment analyses. Larger pathways demonstrated a closer alignment to the identity line, with lower RMSE and higher correlation coefficients, highlighting LDAK-PBAT’s efficiency in handling complex genetic datasets.

### Comparative Performance of LDAK-PBAT with MAGMA and Hypergeometric Testing Across Various Statistical Thresholds

Figure 4 compares the performance of LDAK-PBAT, MAGMA, and Hypergeometric testing across p-value thresholds of 5e-02, 3.125e-03, and 2.78e-06 using sensitivity, precision, F1 score, and specificity. Across p-value thresholds, LDAK-PBAT outperformed MAGMA and Hypergeometric testing in F1 score (0.734 vs. 0.636 and 0.570 at 5e-02) and demonstrated superior precision at the strictest threshold (0.839 vs. 0.961 and 0.761 at 2.78e-06). While MAGMA had the highest specificity (0.862 at 5e-02), its sensitivity sharply declined at lower thresholds (0.117 at 2.78e-06). Hypergeometric testing provided consistent but moderate results across all metrics. These findings underscore LDAK-PBAT’s robust precision in detecting true positives, crucial for efficient genetic pathway analysis.

**Figure 4.**
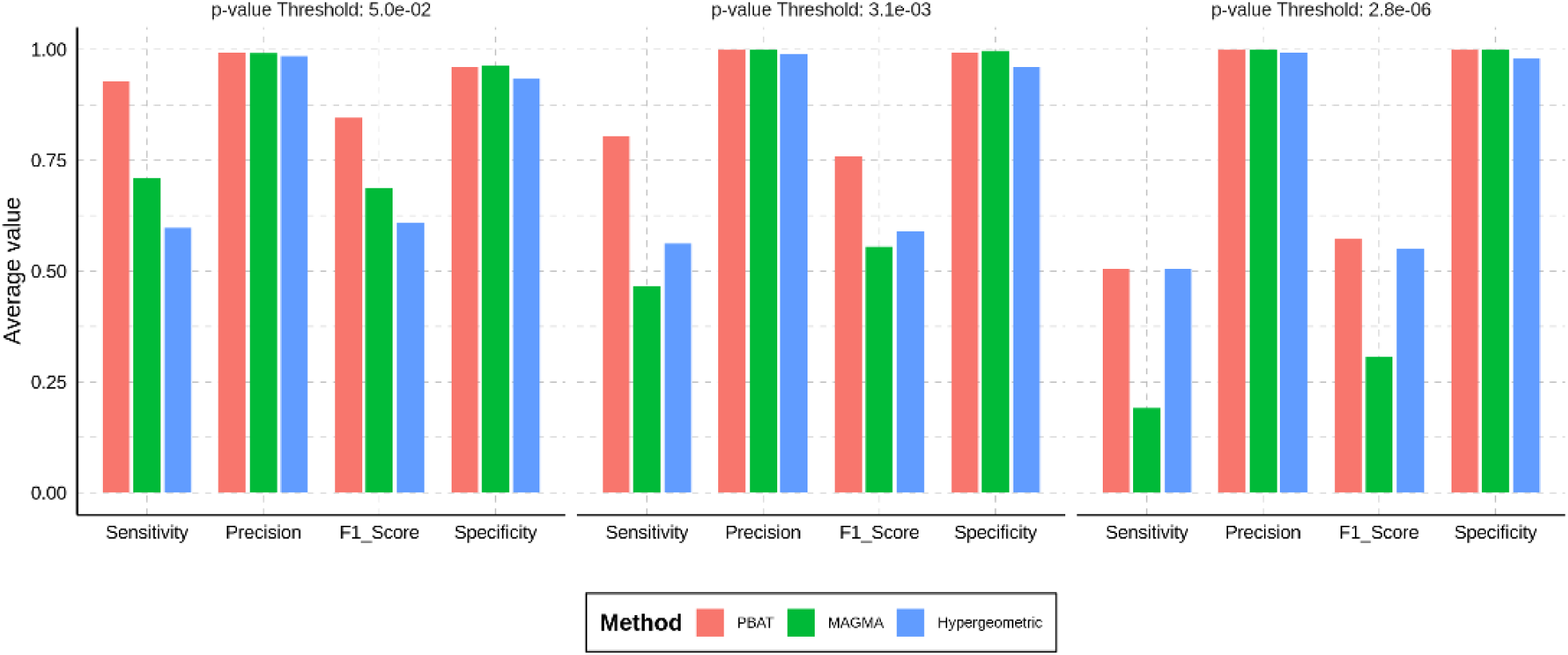
Comparative Analysis of LDAK-PBAT with MAGMA and Hypergeometric Testing Methods Across Multiple Statistical Thresholds. This figure displays a bar chart comparing the performance of LDAK-PBAT with two other methods, MAGMA and Hypergeometric testing, across different p-value thresholds (5e-02, 3.1e-03, and 2.8e-06). Four key metrics are evaluated: sensitivity, precision, F1 score, and specificity. The average values for each metric are shown at each p-value threshold, with separate bars representing each method. The comparison is intended to demonstrate the relative strengths of LDAK-PBAT in identifying true positive associations within genetic datasets.

### Superior Detection of associated Pathways in real Data Using LDAK-PBAT

The analytical depth of Figure 5 underscores the differential detection capabilities of genomic pathway analysis methods using real data from 37 traits from UKBB, PGC and MVP. At a nominal p-value threshold of 0.05, LDAK-PBAT surfaces with 34858 significant pathways, eclipsing the findings of MAGMA, which detects 17704 pathways, and various configurations of Hypergeometric testing, which identify between 7057 and 8882 pathways. This indicates LDAK-PBAT’s pronounced sensitivity in identifying potential genetic associations.

**Figure 5.**
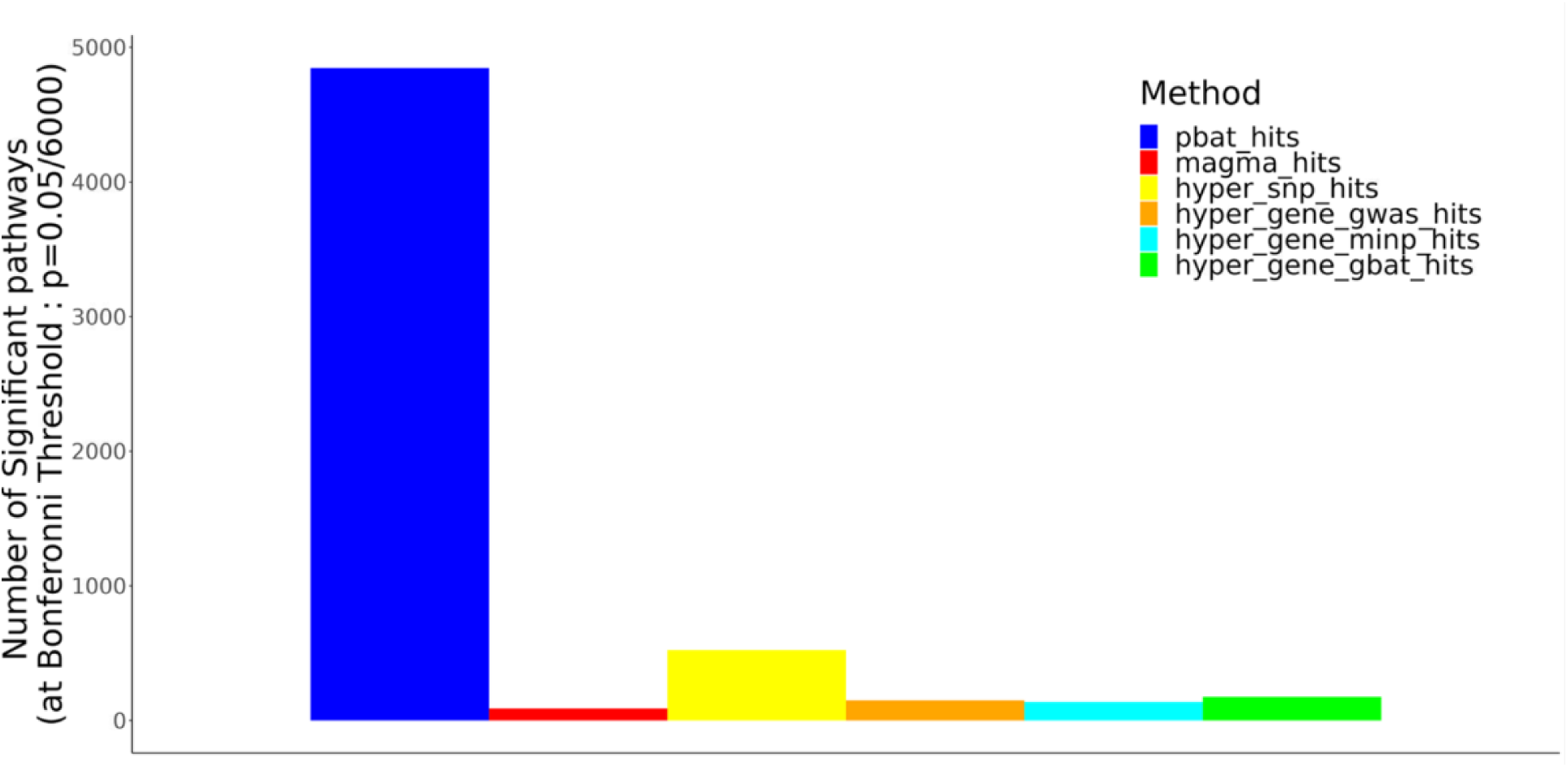
Comparative Efficacy of Pathway Analysis Methods across 37 traits from the UK Biobank, MVP and PGC datasets. The bar plots enumerate the count of significant pathways at the Bonferroni-adjusted threshold (0.05/6000), delineating each method by its number of significant pathways detected. pbat_hits: Pathways identified as significant by LDAK-PBAT, using a heritability-based framework to directly test pathway-level associations. magma_hits: Pathways identified as significant by MAGMA, which uses a two-step approach by first aggregating SNPs to genes and then testing pathways. hyper_snp_hits: Pathways enriched for significant clumped SNPs based on GWAS summary statistics. hyper_gene_gwas_hits: Pathways enriched for genes harboring GWAS-significant SNPs. hyper_gene_minp_hits: Pathways enriched for genes whose minimum SNP p-value passes a Bonferroni threshold based on the total number of genes. hyper_gene_gbat_hits: Pathways enriched for significant genes identified using GBAT, a gene-based association testing method, with a Bonferroni threshold based on the total number of genes.

At the more stringent Bonferroni threshold of 8e-6, LDAK-PBAT’s robustness is further corroborated, with 4861 significant pathways identified, compared to MAGMA’s 97. Hypergeometric testing under this threshold exhibits a limited discovery scope, with the gene-based test (GBAT) uncovering 179 pathways and the SNP clump-based approach revealing 522, while the gene GWAS and minP tests detect 157 and 145 pathways, respectively.

These findings indicate that LDAK-PBAT’s power advantage is not confined to a single trait but is consistently observed across a wide range of traits. For instance, the supplementary table 1 shows that LDAK-PBAT identifies 283 significant pathways for height, 403 for Alzheimer’s disease, and 857 for LDL cholesterol, whereas MAGMA and the Hypergeometric tests identify significantly fewer pathways across the board. This suggests that the superior performance of PBAT is widespread, making it a valuable tool for large-scale genetic analyses and providing enhanced insights into complex traits.

### Validation of Pathway Significance Across Increasing Sample Sizes: A Comparative Study with LDAK-PBAT and Other Methods

In pathways analysis of ten UKBB traits using LDAK-PBAT with an initial sample size of 50,000 individuals, we identified 348 pathways with significant associations. To validate these pathways, we extended the analysis to larger sample sizes and utilized various established methods for comparison. As depicted in Figure 6, of the pathways identified by LDAK-PBAT, 9 (2.6%) were also found significant by other methods combined at the 50k sample size. This number increased as the sample size expanded, with 33 (9.5%) at 100k, 71 (20.4%) at 150k, and 108 (31.0%) at 200k retaining their significance. Notably, the method Hyper_SNP_hits confirmed 101 of these pathways at the largest sample size of 200k, while Hyper_Gene_GBAT_hits identified 19. The consistent increase in validated pathways corroborates the initial findings from LDAK-PBAT, indicating a substantial proportion of these associations are likely to be relevant to the genetic architecture of these traits.

**Figure 6.**
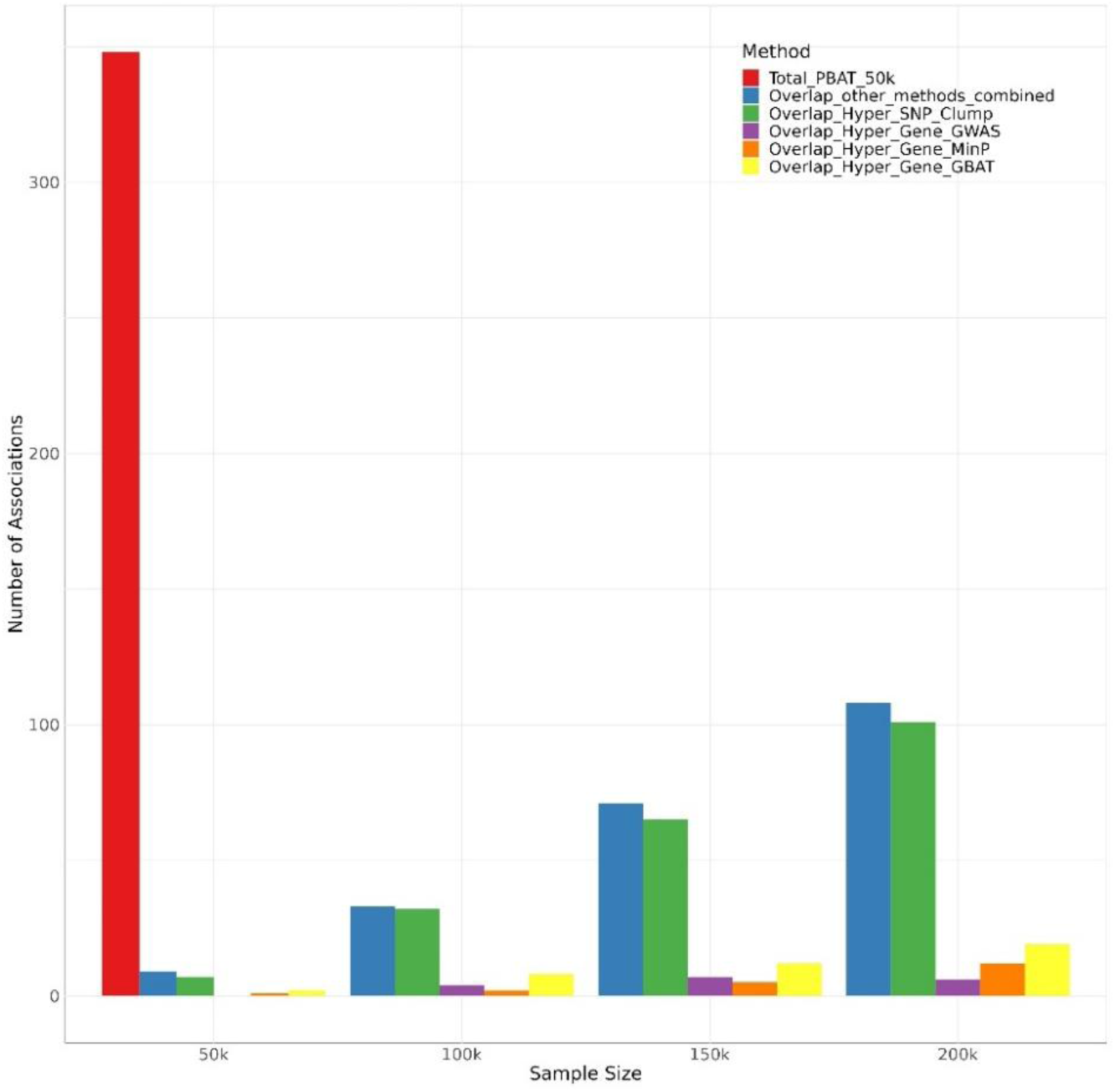
Comparative Analysis of Pathway Significance Across Increasing Sample Sizes Using LDAK-PBAT and Other Methods. The visualization displays the initial set of pathways deemed significant by LDAK-PBAT at a stringent p-value threshold of 0.05/6000, corresponding to the number of analysed pathways. The Red bar represent these PBAT-identified pathways using a 50k sample size. The subsequent bars of varying colours display the count of PBAT-identified pathways that maintain significance when evaluated by other methods (Hyper SNP Clump, Hyper Gene GWAS, Hyper Gene MinP, Hyper Gene GBAT) as sample sizes increase incrementally from 50k to 200k. This progression highlights the robustness and reproducibility of significant associations detected by LDAK-PBAT when subjected to validation by alternative methods with larger datasets.

### Robustness of LDAK-PBAT Against Variations in SNP Lists and Reference Panels

In our sensitivity analysis involving a subset of 225 simulated phenotypes and 100 pathways, we examined the LDAK-PBAT model’s robustness against SNP list and reference panel variations. When comparing models based on 1 million HapMap SNPs to those utilizing an expanded list of 7.1 million SNPs, heritability enrichment showed a high degree of concordance, with a correlation coefficient of 0.784 (Supplementary Figure 1A). The consistency of p-value significance was even more pronounced, evidenced by a correlation of 0.996 (Supplementary Figure 1B). The average difference in heritability enrichment between the two SNP models from UK Biobank was minimal (mean difference = 0.028, SD = 3.866), suggesting a negligible effect of SNP density on the enrichment estimates. A slight negative mean difference in -log10(p-value) between the models (mean difference = -0.292, SD = 0.865) indicates a minor inflation in significance when utilizing the full SNP list. These findings illustrate that increasing the SNP list within the same reference panel maintains the integrity of heritability estimations.

**Supplementary Figure 1.**
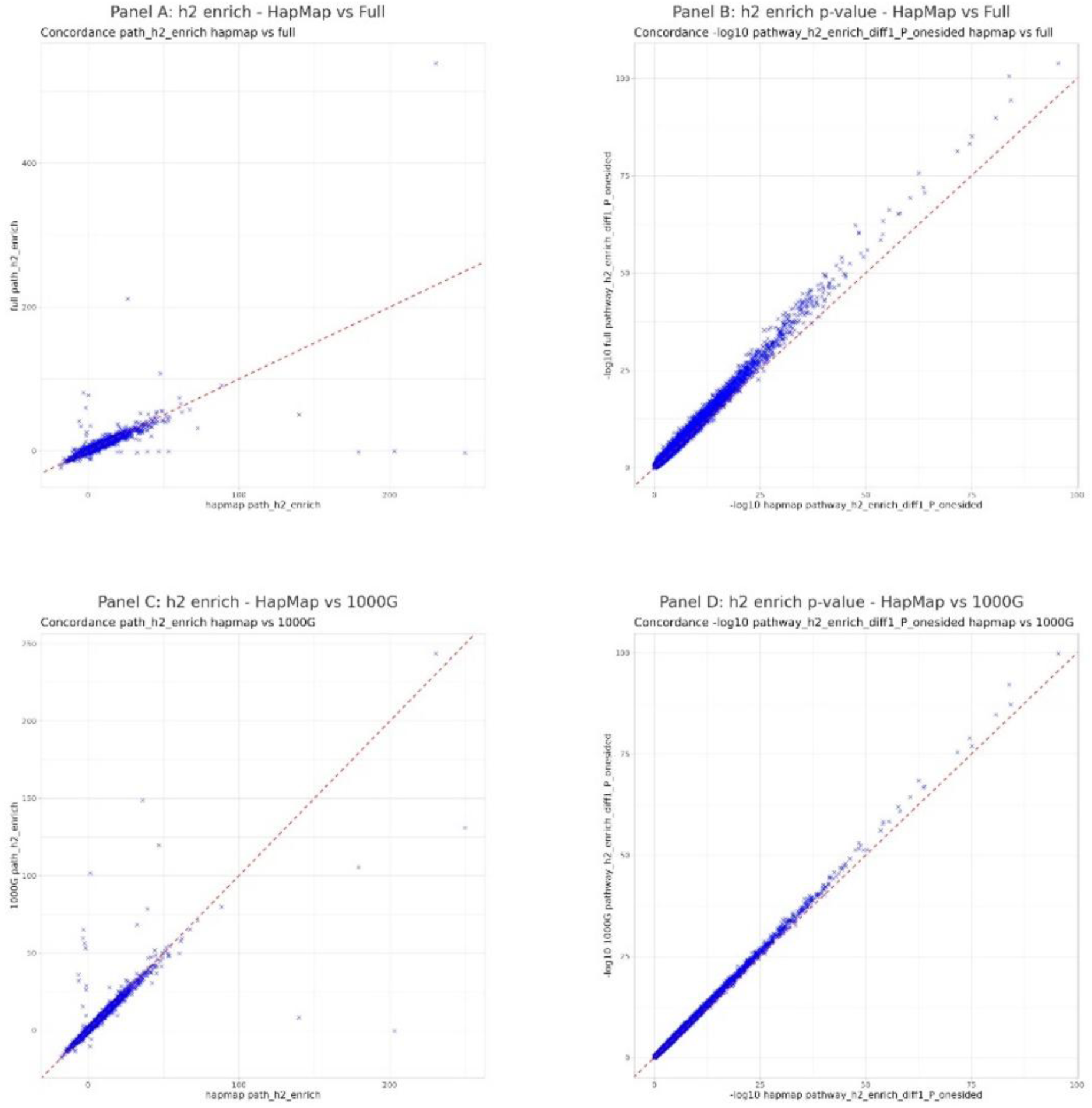
Assessment of LDAK-PBAT Model robustness to SNP List and Reference Panel Variations. Panels A and B illustrate the comparison of heritability enrichment (h2 enrich) and p-value significance between two LDAK-PBAT analyses using the same UK Biobank individuals (n=2500) as reference panel but different SNP list: HapMap (1 million SNPs) versus comprehensive (7.1 million SNPs) list. Panels C and D focus on the comparison of heritability enrichment and p-value significance between LDAK-PBAT analyses using the same HapMap SNP list (1 million SNPs) but different reference panels: UK Biobank individuals (n=2500) versus 404 non-Finnish European individuals from the 1000 Genomes Project (1000G). These panels demonstrate the model’s consistency across variations in SNP selection and reference population sources.

Further investigation into the impact of changing reference panels, while holding the SNP list constant, revealed a correlation in heritability enrichment of 0.901 when comparing UK Biobank to the 1000G non-Finnish European individuals (Supplementary Figure 1C). The p-value significance correlation was near perfect at 0.999 (Supplementary Figure 1D), indicating that the choice of reference panel, even when different, does not influence the statistical significance of heritability enrichment. Comparisons between different reference panels using the HapMap SNP list showed a mean difference in heritability enrichment of 0.026 (SD = 2.442), corroborating the high correlation observed. The difference in -log10(p-value) was marginal (mean difference = - 0.114, SD = 0.302), reinforcing the notion that the reference panel selection has a limited effect on the significance of heritability enrichment findings.

Overall, the LDAK-PBAT model demonstrates considerable resilience to changes in SNP list and reference panel selection. The slight variations detected in heritability enrichment do not undermine the conclusions drawn from the p-value significance, attesting to the reliability of the enrichment analysis approach employed in this study.

### Robustness of LDAK-PBAT to the Exclusion of Genic Annotations

Supplementary Figure 2 compares pathway *Z*_Stat2_ values between the heritability model incorporating the genic component and the model excluding it. A total of 4,753 pathways were consistently identified as significant (*Z*_Stat2_≥4.3054) across both models, demonstrating strong agreement in results and robustness of LDAK-PBAT for most declared significant pathways. However, 96 pathways were significant only when the genic component was included, while 308 pathways were significant only when it was excluded, highlighting the influence of genic SNPs on pathway enrichment under specific genetic architectures. A small subset of pathways (flagged as outliers in Supplementary Figure 2) exhibited highly discordant *Z*_Stat2_ ratios between the two models, suggesting that the exclusion of the genic component leads to inflation in pathway-level *Z*_Stat2_ values in some cases. While this inflation does not alter the overall conclusions for most significant pathways, it supports the inclusion of the genic component in the default model to maintain more accurate pathway enrichment estimates. These results demonstrate that LDAK-PBAT remains robust in identifying significant pathways but performs optimally when the genic component is included.

**Supplementary Figure 2.**
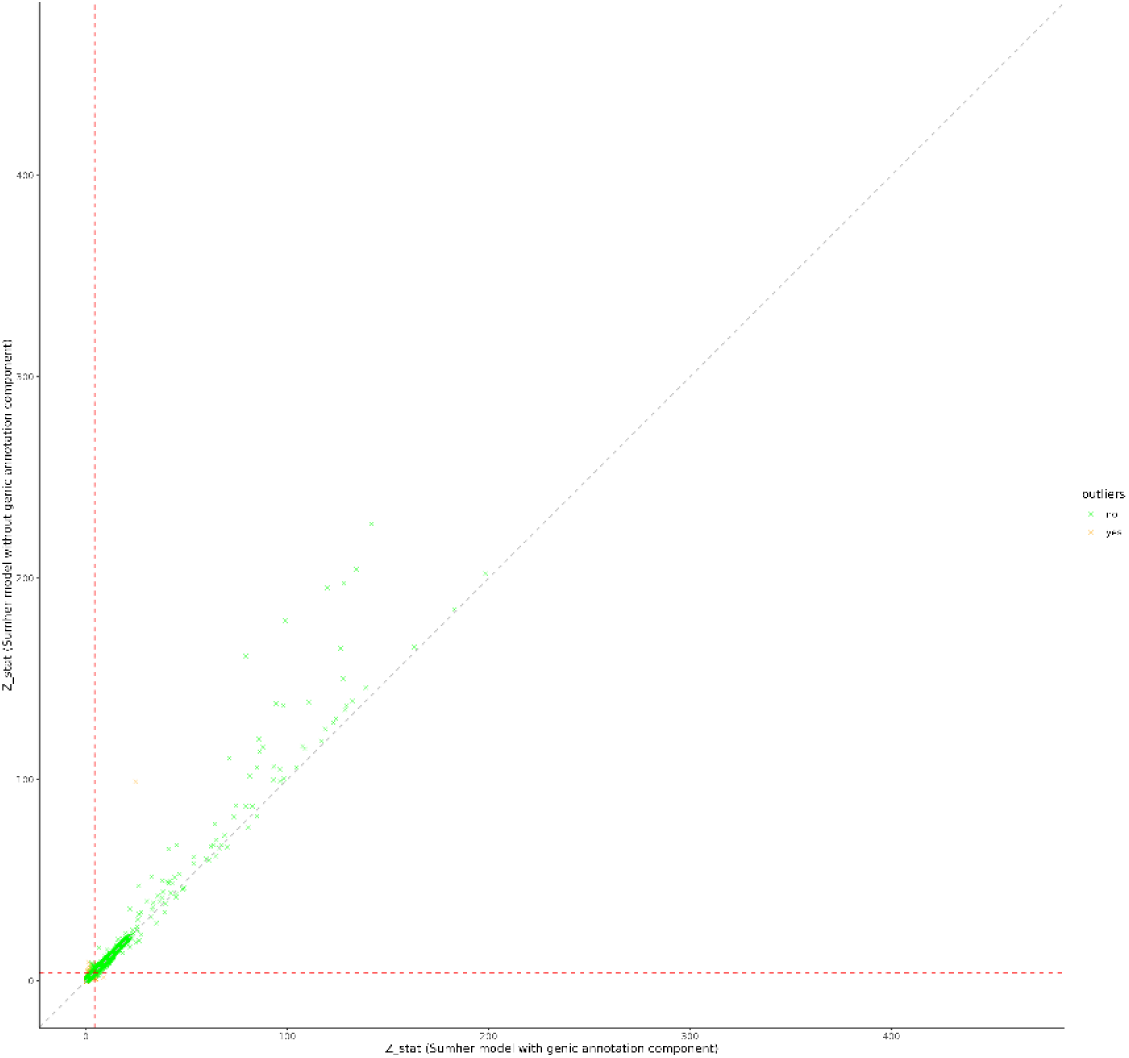
Comparison of Pathway ZZZ-statistics from LDAK-PBAT with and without the genic annotation component. The scatterplot compares pathway Z_Stat2_ values obtained using the full heritability model (including the genic annotation component; x-axis) versus the modified model excluding the genic component (τ_2_; y-axis). Green points represent pathways with consistent enrichment results between models, while orange points highlight outliers with highly discordant Z-statistic ratios (>3 or <1/3). Dashed lines correspond to the Bonferroni threshold (Z=4.3054). The results demonstrate broad consistency between models but identify pathways sensitive to the inclusion of the genic component.

### Adding an Intercept Modestly Alters Pathway Enrichment Results

Supplementary Figure 3 illustrates the comparison of Z_Stat2_ values between the default heritability model and the extended model with the intercept. A total of 4,345 pathways were consistently identified as significant (Z_Stat2_≥4.3054) across both models, indicating broad agreement in results. However, 462 pathways were significant only with the intercept excluded, while 108 pathways were significant only with the intercept included, reflecting sensitivity of pathway enrichment estimates to this adjustment.

**Supplementary Figure 3.**
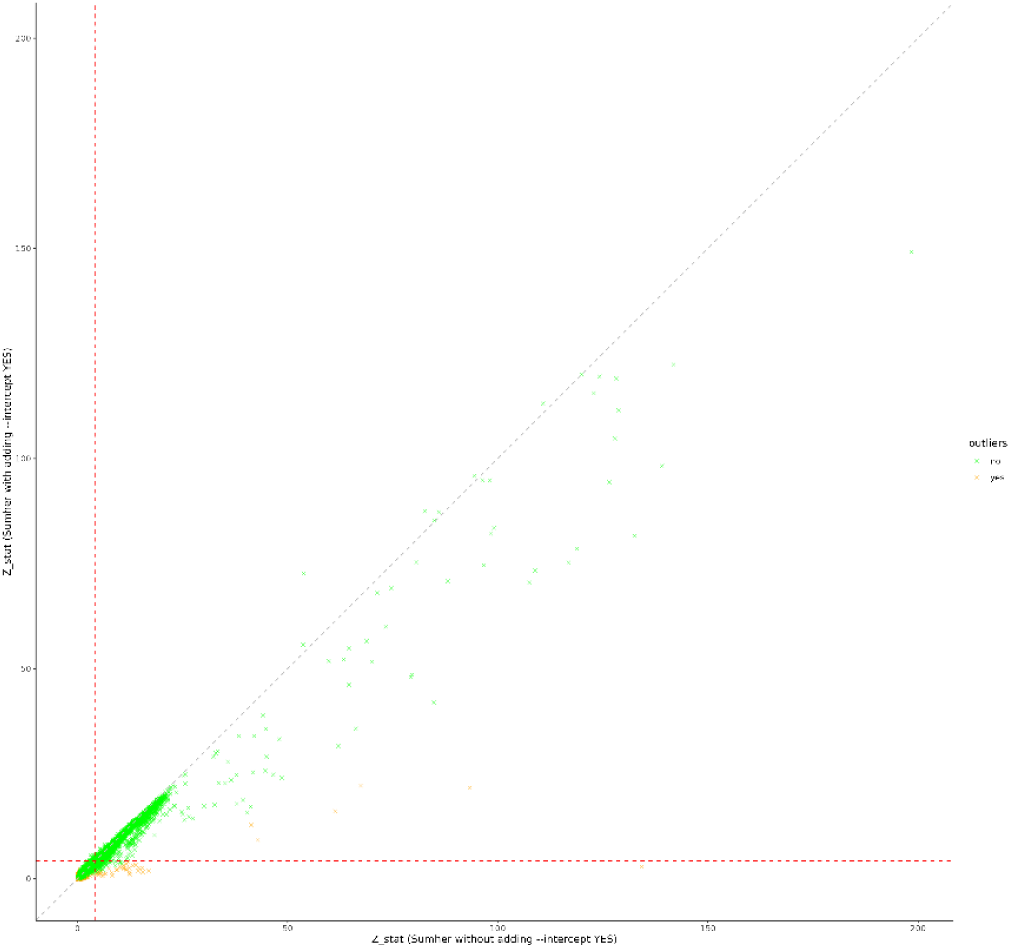
Comparison of Pathway Z_Stat2_ values from LDAK-PBAT with and without the intercept term. The scatterplot compares pathway Z_Stat2_ values obtained using the default heritability model without the intercept term (x-axis) versus the model with the intercept term included (y-axis). Green points represent pathways with concordant enrichment results, while orange points indicate outliers with highly discordant Z_Stat2_ ratios (>3 or <1/3). Dashed lines correspond to the Bonferroni threshold (Z=4.3054).

Adding the intercept reduced the number of significant pathways across all thresholds, particularly at the Bonferroni-adjusted level (0.05/6000), where the number of significant pathways decreased from 4,861 to 4,468. This reduction highlights a potential loss of power when the intercept is included, consistent with its tendency to reduce Z_Stat2_ values. Furthermore, the analysis flagged several outliers (in orange), indicating pathways with highly discordant Z_Stat2_ values between the two models.

These findings demonstrate that adding an intercept term to account for confounding reduces power and alters pathway-level enrichment results. While the default model without the intercept generally performs well, this analysis underscores the importance of carefully considering the assumptions underlying intercept-based adjustments in pathway enrichment studies.

## IV. Discussion

In this study, we introduced and validated LDAK-PBAT, a novel pathway-based analysis tool designed to enhance the detection of genetic associations in complex diseases. Through extensive simulations and analysis of real-world datasets, LDAK-PBAT demonstrated superior performance in estimating pathway heritability and detecting significant pathways compared to existing methods.

The validation of LDAK-PBAT through simulations involving 1,125 continuous phenotypes and 2,250 binary phenotypes confirmed its robustness and accuracy. By varying key genetic parameters such as heritability levels (0.1, 0.3, and 0.5), the number of causal SNPs (2,000, 5,000, and 10,000), and annotation enrichment weights, we created realistic genetic architectures to test the tool’s performance. The results, illustrated in Figures 2 and 3, showed that LDAK-PBAT provides precise heritability estimates and strong correlations with true values across different scenarios. This highlights the tool’s ability to handle complex genetic datasets effectively.

When benchmarked against established gene-set analysis methods like MAGMA and hypergeometric testing, LDAK-PBAT consistently outperformed these tools in terms of sensitivity, precision, and F1 score. As shown in Figure 4, LDAK-PBAT detected a higher number of significant pathways across various statistical thresholds, underscoring its enhanced capability in identifying true genetic associations. This superior performance is particularly notable at stringent thresholds, where maintaining high precision and sensitivity is crucial.

The conceptual differences between PBAT and other methods further explain its superior performance. PBAT has a distinct advantage over hypergeometric gene based due to its ability to account for the magnitude of effects, making it particularly powerful when a pathway contains many small-effect genes, most of which are not individually significant. In contrast to MAGMA, LDAK-PBAT’s strength lies in its approach to considering all pathway SNPs together, rather than first dividing them into genes. Then using an approximate method to correct for the overlap and LD between genes. Additionally, while PBAT is like hypergeometric SNP based, it accounts for linkage disequilibrium (LD) and employs a heritability model, which further enhances its effectiveness in detecting significant pathways.

The application of LDAK-PBAT to 37 traits from the UK Biobank, MVP, and PGC datasets further demonstrated its efficacy. Figure 5 highlights the tool’s ability to detect significantly more pathways at nominal and Bonferroni-adjusted thresholds compared to other methods. This comprehensive analysis confirms that LDAK-PBAT can provide valuable insights into the genetic basis of complex diseases, making it a potent tool for large-scale genetic studies.

The robustness of LDAK-PBAT was further assessed through sensitivity analyses involving variations in SNP lists and reference panels. As depicted in Supplementary Figure 1, the tool maintained high concordance in heritability enrichment estimates and p-value significance despite changes in SNP density and reference population. This consistency reinforces the reliability of LDAK-PBAT’s enrichment analysis approach, ensuring that the tool’s conclusions are not significantly affected by these variations.

Despite its strengths, LDAK-PBAT has certain limitations that should be acknowledged. Firstly, the tool’s performance is dependent on the quality and completeness of pathway definitions from databases like MSigDB. Inaccuracies or biases in these definitions could affect the results. Secondly, while LDAK-PBAT handles a broad range of genetic architectures, extremely rare variants or small effect sizes might still pose challenges for accurate detection. Additionally, the initial step of constructing the model for LDAK-PBAT can be computationally demanding. However, this model is built once and then provided for subsequent analyses, significantly reducing the computational burden for users who apply the pre-built model with summary statistics to analyze pathway associations. This makes the tool more accessible and efficient for large-scale genetic studies.

LDAK-PBAT’s ability to accurately estimate pathway heritability and detect significant pathways has important implications for genetic research and personalized medicine. By focusing on the collective impact of multiple genes within biological pathways, LDAK-PBAT offers a more holistic view of genetic influences on complex traits. This approach can lead to the identification of novel therapeutic targets and a better understanding of disease mechanisms.

Future work will involve expanding the tool’s capabilities to include additional datasets and refining its algorithms to further enhance its performance. Additionally, integrating functional annotations and epigenetic data could provide deeper insights into the regulatory mechanisms underlying complex traits.

In conclusion, LDAK-PBAT is a robust and reliable tool for pathway-based genetic analysis, offering significant advantages over existing methods. Its precise estimation of pathway heritability, superior detection of significant pathways, and resilience to variations in SNP lists and reference panels make it an asset for genetic researchers. LDAK-PBAT is freely available within the LDAK software package, providing researchers with the resources needed to apply this powerful tool to their own genetic studies.

## Data and code availability

Instructions for running LDAK-PBAT are available on the LDAK website (https://www.ldak.org), while sample code for the analyses in this paper is on the GitHub PAGE of T.-E.B. (https://github.com/takiy-berrandou/LDAK-PBAT-paper-scripts). We applied for and downloaded individual-level UK Biobank data from the UK Biobank website (https://www.ukbiobank.ac.uk). We applied for and downloaded MVP summary statistics from the dbGaP website (https://www.ncbi.nlm.nih.gov/projects/gap/cgi-bin/study.cgi?study_id=phs001672.v3.p1). We downloaded PGC summary statistics from the PGC website (https://www.med.unc.edu/pgc/download-results).

## Acknowledgments

T.-E. B. is supported by FRM grant (ARF202309017669). D.S. is supported by a European Research Council Consolidator Grant (ID 101088901, acronym ClassifyDiseases)

## Author contributions

Writing and editing the manuscript: T.-E.B. and D.S. Study design/conception: T-E.B., and D.S. Analyses: T.-E.B. and D.S.

## Declaration of interests

The authors declare no competing interests.

## Supplemental information

Supplementary table 1: Comparative Efficacy of Pathway Analysis Methods for each 37 traits from the UK Biobank, MVP and PGC datasets.

## Web resources

LDAK-PBAT, https://www.ldak.org

MAGMA, https://ctg.cncr.nl/software/magma

Million Veteran Program summary statistics, https://www.ncbi.nlm.nih.gov/projects/gap/cgi-bin/study.cgi?study_id=phs001672.v3.p1

Psychiatric Genomics Consortium summary statistics, https://www.med.unc.edu/pgc/download-results

T-E.B.’s GitHub, https://github.com/takiy-berrandou/LDAK-PBAT-paper-scripts

UK Biobank, https://www.ukbiobank.ac.uk

## Notes

### Competing Interest Statement

The authors have declared no competing interest.

